# Protection of previous SARS-CoV-2 infection is similar to that of BNT162b2 vaccine protection: A three-month nationwide experience from Israel

**DOI:** 10.1101/2021.04.20.21255670

**Authors:** Yair Goldberg, Micha Mandel, Yonatan Woodbridge, Ronen Fluss, Ilya Novikov, Rami Yaari, Arnona Ziv, Laurence Freedman, Amit Huppert

**Affiliations:** Technion - Israel Institute of Technology, Israel; The Hebrew University of Jerusalem, Israel; The Gertner Institute for Epidemiology & Health Policy Research, Sheba Medical Center, Israel; Tel Aviv University, Israel

**Keywords:** vaccine efficacy, COVID-19, SARS-CoV-2, previous infection, protection from reinfection

## Abstract

Worldwide shortage of vaccination against SARS-CoV-2 infection while the pandemic is still uncontrolled leads many states to the dilemma whether or not to vaccinate previously infected persons. Understanding the level of protection of previous infection compared to that of vaccination is critical for policy making. We analyze an updated individual-level database of the entire population of Israel to assess the protection efficacy of both prior infection and vaccination in preventing subsequent SARS-CoV-2 infection, hospitalization with COVID-19, severe disease, and death due to COVID-19. Vaccination was highly effective with overall estimated efficacy for documented infection of 92·8% (CI:[92·6, 93·0]); hospitalization 94·2% (CI:[93·6, 94·7]); severe illness 94·4% (CI:[93·6, 95·0]); and death 93·7% (CI:[92·5, 94·7]). Similarly, the overall estimated level of protection from prior SARS-CoV-2 infection for documented infection is 94·8% (CI:[94·4, 95·1]); hospitalization 94·1% (CI:[91·9, 95·7]); and severe illness 96·4% (CI:[92·5, 98·3]). Our results question the need to vaccinate previously-infected individuals.

## Introduction

Israel is currently in the later stages of a vaccination campaign to reduce both SARS-CoV-2 infection and the number of COVID-19 cases. Israel is administering the BNT162b2 vaccine, developed by BioNTech in cooperation with Pfizer,^1^ for which an Emergency Use Authorization (EUA) was issued by the Food and Drug Administration (FDA).^2^ The vaccine is administered in two doses, with a 21-day interval between doses. Israel launched its COVID-19 vaccination program on December 20, 2020. The vaccine became available, free of charge, to different risk groups in stages: first to those older than 60 years old, nursing home residents, healthcare workers, and patients with severe comorbidities, and then gradually to younger age groups. As of February 6, 2021, the vaccine was made available to all individuals aged 16 or older not previously infected by SARS-CoV-2. As of March 20, 2021, 77% of the eligible population is vaccinated. Due to the high caseload and the local detection of viral mutants such as B.1.1.7, Israel went into a third nationwide lockdown during the vaccination campaign. A light lockdown began on December 24, 2020, and was tightened on January 5, 2021. Restrictions were eased in stages starting February 7, 2021. The dynamics of the epidemic as well as the vaccination campaign appear in Figure 1.

**Figure 1:**
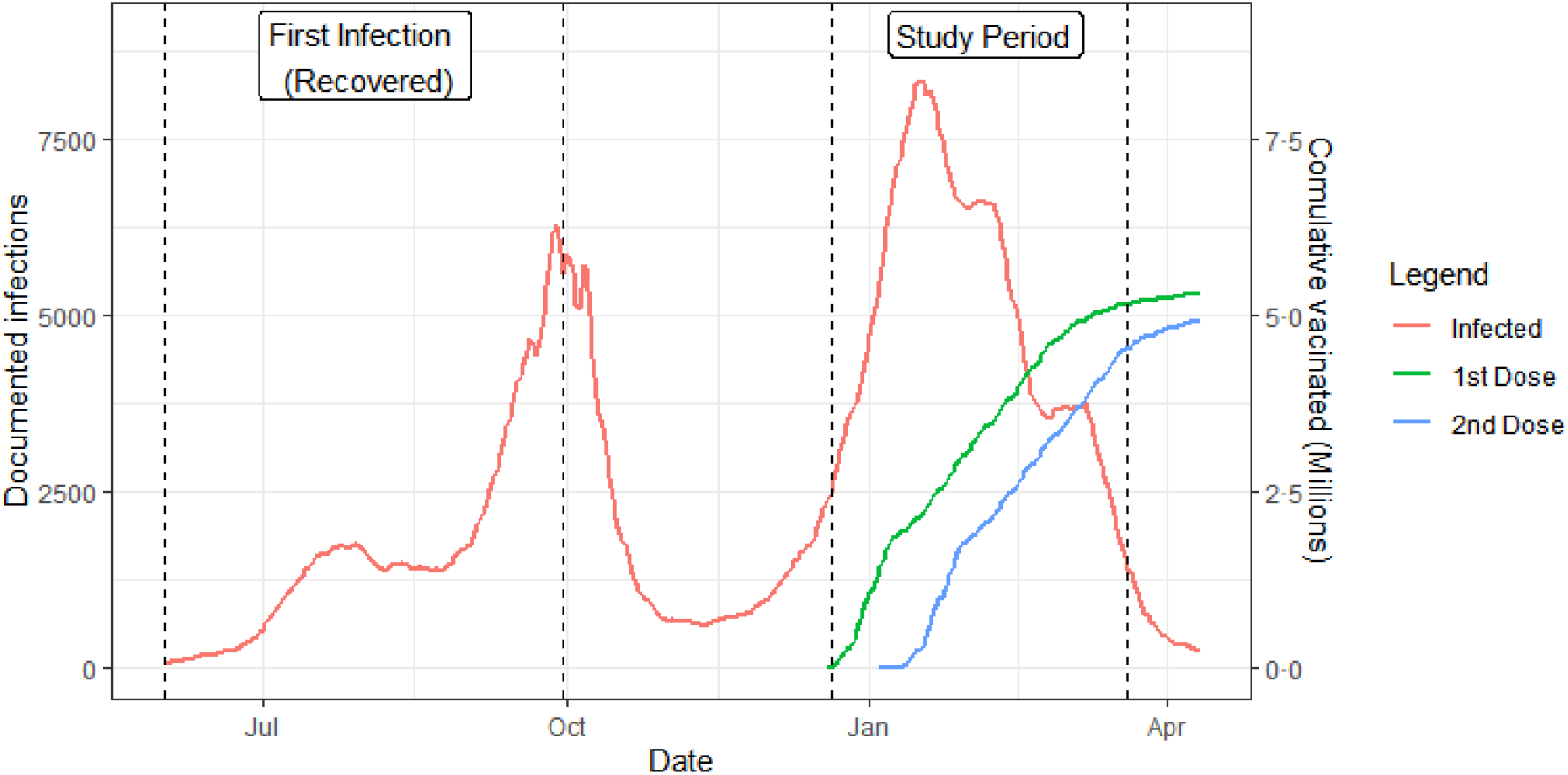
Population dynamics. Documented new infections and cumulative vaccinated persons by date. The study period and the infection period of the recovered cohorts are marked by vertical lines.

SARS-CoV-2 testing in Israel is carried out according to the following policy: individuals may request testing due to either symptoms or contact with an individual who tested positive. These PCR tests are given free of charge. Individuals who have come into contact with an individual who tested positive are required to self-quarantine for 14 days. This quarantine period may be shortened to 10 days if the individual is tested twice during the first 10 days, and both test results are negative. Individuals who have received both vaccine doses, and had the second dose seven days or more before a contact with a positive individual, and do not have symptoms, are not required to self-quarantine, and thus have less motivation to get tested. In addition to voluntary testing, Israel conducts routine testing of all nursing-home workers.

Recent results based on aggregated data^3–5^ and individual level data^6–10^ have shown that the vaccine substantially reduces the number of severe COVID-19 cases. Two studies also indicate that the viral load of vaccinated individuals is significantly reduced.^11,12^ These encouraging initial results are based on a short follow-up of vaccinated individuals. Results on previous COVID-19 infection^13–16^ suggest protection against reinfection compared to uninfected unvaccinated individuals.

In this study, we estimate the efficacy of the vaccine in the reduction of documented SARS-CoV-2 infection and severe COVID-19 disease. We focus on four cohorts: unvaccinated individuals; vaccinated individuals followed from first dose to a week after the second dose; vaccinated individuals followed from a week after the second dose onwards, and the Recovered Cohort of unvaccinated individuals previously infected with SARS-CoV-2. For more details, see the Methods section. All efficacies, of vaccine or previous infection, are compared to the unvaccinated cohort.

The prospective observational analysis that we present faced several challenges. The first challenge was self-selection of treatment, which implies differences in potential risk factors between vaccinated and non-vaccinated individuals. These include age, sex, socio-demographic level,^17^ level of infection in the immediate environment, and possibly other behavioral variables that could affect level of exposure to the virus. The second challenge was detection bias: willingness to undergo vaccination can be associated with trust in the healthcare system, which may also imply a tendency to comply with testing regulations. On the other hand, vaccinated individuals may feel more protected and may ignore mild symptoms indicative of the disease, and have less motivation to get tested as they are not required to self-quarantine after a contact with a positive individual. The third challenge was the variation in infection risk throughout the vaccination campaign, mainly due to varying lockdown levels, relative prevalence of viral mutants, and local outbreaks. Lastly, the status of individuals (i.e., unvaccinated, partially vaccinated, or fully vaccinated) was dynamic: with time, individuals move from one cohort to another, and between risk groups. In the Methods Section we explain how we designed the analysis to address these challenges.

## Methods

### Data

The database included two main tables. The first table was of all 1373 municipalities in Israel, with data on the number of residents, the daily count of PCR tests, and the daily positive results. This table was constructed based on data from the Israel Ministry of Health and the Israel Central Bureau of Statistics.

The second table was an individual-level table on persons aged 16 and above collected by the Israeli Ministry of Health based on data received routinely from all HMOs and hospitals and linked using the person’s identity number. This table contained basic demographic data and information on dates of first and second vaccinations, if received, and dates and results of all PCR tests performed from March 1, 2020, up to March 20, 2021. For individuals with a positive PCR test, the table contained information on symptoms, as well as the maximum severity status throughout the course of the disease (hospitalization, severe disease, death). The definition of hospitalization, severe disease, and death due to COVID-19 is based on international recommendations.^18^ Specifically, hospitalization is defined as being admitted due to COVID-19. Disease is considered severe when a patient has >30 breaths per minute, oxygen saturation on room air <94%, or ratio of arterial 148 partial pressure of oxygen to fraction of inspired oxygen <300mm mercury. Data on symptoms were also available but we found them less reliable and thus did not include symptomatic COVID-19 as an outcome.

Thus, the table contained an entry for every adult (age ≥ 16) in Israel who had at least one PCR test or had received at least the first dose of the vaccine (with a total of 5,682,928 entries). Adults with no PCR test and no vaccination (668,975) were added to the table using data from the Israel Central Bureau of Statistics. Thus, this second table included 6,351,903 entries with basic demographic data of the total adult population in Israel, as well as their PCR tests and vaccination dates. Individuals under age 16 are not eligible for vaccination and were excluded from this study. A summary of the data appears in Table 1.

**Table 1:** Population level data. Columns Male, Female, and Total are in thousands. Columns PCR tests, Positive tests, Hospitalized, Severe, and Death, are the counts during the period December 20, 2020 to March 20, 2021.

| Age | Male | Female | Total | PCR | Positive | Hospitalization | Severe | Death |
| --- | --- | --- | --- | --- | --- | --- | --- | --- |
| 16-39 | 1,513 | 1,484 | 2,997 | 2,414,803 | 183,617 | 2,722 | 684 | 44 |
| 40-49 | 531 | 542 | 1,073 | 810,988 | 49,373 | 1,614 | 814 | 64 |
| 50-59 | 404 | 423 | 827 | 575,853 | 34,411 | 1,978 | 1,252 | 153 |
| 60-69 | 345 | 386 | 731 | 399,149 | 21,073 | 2,242 | 1,528 | 406 |
| 70-79 | 207 | 249 | 456 | 207,538 | 10,410 | 2,358 | 1,757 | 674 |
| 80+ | 106 | 161 | 267 | 197,916 | 7,828 | 3,105 | 2,428 | 1,386 |
| Total | 3,107 | 3,245 | 6,352 | 4,606,247 | 306,712 | 14,019 | 8,463 | 2,727 |

To account for environmental risk, we calculated a municipality daily risk index by the number of cases newly confirmed in the past seven days per 10,000 residents. We used a 7-day moving average since the number of PCR tests typically drops at weekends. The index was categorized into four risk levels (up to one, one to four, four to ten, and more than ten daily cases per 10,000) to yield the municipality daily risk category, and was used as a covariate in the risk model.

Behavioral differences among people may result in different levels of exposure to infection and compliance with PCR testing guidelines. We partially accounted for this by counting the number of PCR-test clusters that an individual underwent from March 1, 2020, to December 20, 2020 (i.e., prior to the vaccination program). Here, a PCR-test cluster comprised all consecutive test performed within 10 days of each other. We then defined three individualized background risk levels: no PCR tests, one cluster, and two or more clusters, and this covariate was also included in the risk model. For previously-infected individuals, we set the level to one cluster and checked sensitivity to this value. Note that the time interval for defining this variable (up to December 20, 2020) did not overlap with the follow-up period.

In addition to estimating vaccine efficacy, we estimated the protection of prior SARS-CoV-2 infection against a recurrent infection. Thus, we also included in the dataset individuals who had recovered from COVID-19. Recovery from SARS-CoV-2 infection is not well-defined, and individuals may continue to show traces of the virus weeks and sometimes even months after the infection.^14^ We defined as a recurrent infection only cases occurring three months or more after the first diagnosis. We also considered only individuals for whom the first infection was diagnosed between June 1 and September 30, 2020, as the PCR results before June 1 are considered less reliable. Hence, individuals infected before June 1, 2020 or between October 1, 2020 and December 20, 2020 were excluded from the analysis.

### Statistical Modeling

To estimate the efficacy of the Pfizer BNT162b2 vaccine in reducing documented SARS-CoV-2 infection and other COVID-19 events, we considered four dynamic sub-populations or cohorts:

- Cohort 0: Unvaccinated and not previously infected with SARS-CoV-2;
- Cohort 1: Vaccinated and followed from the day of first vaccination to 6 days after the second dose;
- Cohort 2: Vaccinated and followed from a week after the second dose onwards;
- Recovered: Unvaccinated and previously diagnosed with SARS-CoV-2 between June 1 and September 30, 2020.

On any given calendar day, each individual included in the analysis belongs to a single cohort, but cohort membership is dynamic. Moreover, individuals may not only move between cohorts over time (for example, from cohort 0 to cohort 1 after first vaccination, or from cohort 1 to cohort 2 at 7 days after the second vaccination), but also exit from the follow-up (for example, on infection with SARS-CoV-2 or death). The outcomes hospitalization, severe disease, and death, were attributed to the date on which COVID-19 was documented.

We modeled the daily risk of each individual from December 20, 2020 to March 20, 2021, as a function of calendar time, the cohort to which the individual currently belonged, and the individual’s current risk factors, which included fixed covariates: age group (16-39, 40-49, 50-59, 60-69, 70-79, and 80+), sex, and background risk level (0,1, and 2+ past PCR tests), and the time-dependent variable: municipality risk level(low, medium, medium-high, and high). We refer to each combination of possible covariate values (age group, sex, background risk level, and municipality risk level) as the risk profile.

Our analysis model falls within the framework of multi-state survival models, where each cohort represents a separate state;^19^ see Figure S1. Similar to the study of mRNA-1273, the vaccine developed by Moderna,^20^ we defined the efficacy of the vaccine in terms of hazard ratios, where the main interest is in comparing the hazard of a non-vaccinated individual (Cohort 0) to that of an individual who had completed the recommended protocol (Cohort 2). Hazard ratios between cohorts and for each adjusting covariate were estimated via a generalized linear model with a Poisson distribution and logarithmic link function, and an offset for each risk profile.^21^

Our model assumes that for a given cohort and risk profile, the hazard was constant and did not depend on the time from the second dose (Cohort 2). Obviously, the hazard of individuals who have never received the first dose (Cohort 0) cannot depend on the time of the first dose, but we also assumed that the time elapsed from the second vaccination did not affect the hazard in Cohort 2. In other words, we assumed that the protection level did not change with time after the “completion” of the vaccination protocol. While protection by vaccination is expected to decrease in the long run, our assumption is reasonable given the time frame of only three months after first vaccination, where waning immunity is not expected to play a role. We split Cohort 1 into two sub-cohorts: Cohort 1A from the first dose to two weeks after the first dose, and Cohort 1B from 15 days after the first dose to six days after the second dose. Following Skowronski and De Serres,^22^ we considered, as a crude approximation, a constant hazard for each of these two sub-cohorts for every risk profile. To estimate the level of protection among the Recovered Cohort, we made a similar assumption, that the time elapsed from SARS-CoV-2 infection did not affect the hazard ratio.

The formal definition of vaccine efficacy adopted was as follows. Consider any particular risk profile. Let *h*_*i*_ denote the hazard of an individual in one of the vaccinated cohorts 1A, 1B, 2, or Recovered, and let *h*_0_ be the hazard of an individual having the identical risk profile in the unvaccinated group. Efficacy of the vaccine in that cohort for that risk profile is defined as 1 − *h*_*i*_/*h*_0_. Note that the calendar time affects the hazards of the different cohorts only via the time-dependence of the municipality risk level. From the model assumptions, the ratio *h*_*i*_/*h*_0_ is the same for each risk profile, so the estimate of vaccine efficacy may be combined over all the risk profiles. For more details about the model, see Appendix. We analyzed efficacy separately for each of the following outcomes: documented infection, hospitalization, severe disease, and death.

## Results

The data are based on follow-up of the four cohorts from December 20, 2020 up to March 20, 2021, with over 573 million person-days of follow-up. The lengths of follow-up for the fully vaccinated and the recovered cohorts appear in Figures S2 and S3, respectively. During this time 4,606,247 PCR tests were performed (8,040 per million person-days), and 306,712 individuals tested positive (5·4 infections per 10,000 person-days). Of those testing positive, 14,019 (4·6%) required hospitalization, 8,463 (2·8%) were defined as severe cases, and 2,727 (0·9%) died. Table 2 presents these numbers by cohort and age group. The numbers of PCR tests performed per million person-days appear in Table 3. There is a decrease in the rate of PCR testing in both Cohort 2 and the Recovered Cohort compared to the other cohorts. This is likely since fully vaccinated or recovered individuals (Cohorts 2 & Recovered) are more protected against SARS-CoV-2 infection. Additionally, people in Israel need to self-quarantine for 14 days after contacting SARS-CoV-2 infected persons, which can be shortened to ten days if they present two negative PCR tests. This is not required for fully vaccinated and recovered persons unless they develop symptoms.

**Table 2:** Person-day event counts. Person-day counts and event counts for the different cohorts during the period December 20, 2020 to March 20, 2021. Person-day counts are in millions. PCR, Positive, Hospitalized, Severe, and Death, are the actual counts.

| Cohort | Age | Person Days | PCR | Positive | Hospitalization | Severe | Death |
| --- | --- | --- | --- | --- | --- | --- | --- |
| 0 | 16-39 | 170.5 | 1,609,352 | 156,104 | 2,413 | 602 | 38 |
| 0 | 40-49 | 49.4 | 449,371 | 37,075 | 1,331 | 683 | 56 |
| 0 | 50-59 | 31.3 | 268,892 | 23,383 | 1,541 | 1,011 | 122 |
| 0 | 60-69 | 20.5 | 143,320 | 12,130 | 1,528 | 1,051 | 261 |
| 0 | 70-79 | 9.7 | 70,430 | 5,483 | 1,455 | 1,116 | 431 |
| 0 | 80+ | 7.1 | 64,035 | 3,908 | 1,789 | 1,425 | 841 |
| 1A | 16-39 | 27.3 | 287,539 | 19,707 | 231 | 63 | 5 |
| 1A | 40-49 | 11.4 | 107,441 | 7,619 | 201 | 99 | 6 |
| 1A | 50-59 | 9.6 | 85,134 | 6,355 | 290 | 165 | 17 |
| 1A | 60-69 | 8.8 | 61,433 | 4,638 | 400 | 269 | 74 |
| 1A | 70-79 | 6.5 | 30,853 | 2,247 | 418 | 304 | 113 |
| 1A | 80+ | 3.6 | 32,731 | 1,759 | 643 | 490 | 262 |
| 1B | 16-39 | 25.5 | 265,444 | 6,185 | 54 | 11 | 1 |
| 1B | 40-49 | 11.2 | 103,730 | 3,651 | 52 | 20 | 2 |
| 1B | 50-59 | 9.6 | 84,936 | 3,655 | 96 | 52 | 11 |
| 1B | 60-69 | 9.0 | 64,055 | 3,238 | 240 | 160 | 52 |
| 1B | 70-79 | 6.7 | 32,475 | 1,904 | 339 | 244 | 94 |
| 1B | 80+ | 3.7 | 32,244 | 1,440 | 467 | 363 | 204 |
| 2 | 16-39 | 32.9 | 224,106 | 1,002 | 12 | 2 | 0 |
| 2 | 40-49 | 21.7 | 142,540 | 903 | 26 | 12 | 0 |
| 2 | 50-59 | 22.5 | 130,718 | 931 | 44 | 21 | 3 |
| 2 | 60-69 | 27.0 | 126,381 | 1,030 | 69 | 45 | 19 |
| 2 | 70-79 | 21.3 | 72,091 | 764 | 140 | 92 | 36 |
| 2 | 80+ | 11.4 | 67,345 | 707 | 202 | 147 | 78 |
| Recovered | 16-39 | 9.0 | 28,362 | 619 | 12 | 6 | 0 |
| Recovered | 40-49 | 2.4 | 7,906 | 125 | 4 | 0 | 0 |
| Recovered | 50-59 | 1.8 | 6,173 | 87 | 7 | 3 | 0 |
| Recovered | 60-69 | 1.1 | 3,960 | 37 | 5 | 3 | 0 |
| Recovered | 70-79 | 0.5 | 1,689 | 12 | 6 | 1 | 0 |
| Recovered | 80+ | 0.2 | 1,561 | 14 | 4 | 3 | 1 |

**Table 3:** PCR tests per million person days.

| Cohort | 16-39 | 40-49 | 50-59 | 60-69 | 70-79 | 80+ |
| --- | --- | --- | --- | --- | --- | --- |
| 0 | 9,439 | 9,097 | 8,591 | 6,991 | 7,261 | 9,019 |
| 1A | 10,533 | 9,425 | 8,868 | 6,981 | 4,747 | 9,092 |
| 1B | 10,410 | 9,262 | 8,848 | 7,117 | 4,847 | 8,715 |
| 2 | 6,812 | 6,569 | 5,810 | 4,681 | 3,385 | 5,908 |
| Recovered | 3,151 | 3,294 | 3,429 | 3,600 | 3,378 | 7,805 |

We first investigated the dynamics of the vaccination program, disease outcomes, PCR testing, and municipality risk as a function of calendar time. Figures S4 and S5 present the proportion of vaccinated over time among different age and municipality risk groups, respectively. As can be seen from Figure S4, the Israeli vaccination policy was initially to immunize the older population, and as time progressed, younger age groups. Figure S5 shows the association between environmental risk and vaccination. Figure S6 shows the rates over time of the different age groups among those tested, infected, hospitalized, having severe disease, and dying. Table 4 shows, by age group, the estimated vaccine efficacy for the main outcomes for Cohort 2 (fully vaccinated) adjusted for sex, municipality risk, and past PCR. Note that for age groups below 60 years, there were, fortunately, none or very few events of severe illness and death, and thus estimates were omitted for these groups. The table shows that vaccine efficacy was quite similar in all age groups with some decrease in efficacy for the 80+ age category. Fitting a model without age-group/cohort interaction yielded overall vaccine efficacy for documented infection of 92·8% (CI: [92·6, 93·0]); hospitalization 94·2% (CI: [93·6, 94·7]); severe illness 94·4% (CI: [93·6, 95·0]); and death 93·7% (CI: [92·5, 94·7]). We repeated the analysis with full vaccination defined as 15 days or more after the second dose. The results are similar (not shown).

**Table 4:** Vaccination efficacy. Vaccination efficacy for the different age groups adjusted for sex, municipality risk, and past PCR. The overall estimates are based on models without cohort-age interaction. Estimates are not provided for Severe and Death outcomes for the lowest age groups due to very low case numbers in the vaccinated cohorts.

| Age | Positive | Hospitalized | Severe | Death |
| --- | --- | --- | --- | --- |
| 16-39 | 95.1% [94.8, 95.4] | 96.5% [93.8, 98.0] | — | — |
| 40-49 | 92.5% [92.0, 93.0] | 94.4% [91.7, 96.2] | — | — |
| 50-59 | 92.7% [92.2, 93.1] | 95.0% [93.3, 96.3] | — | — |
| 60-69 | 92.4% [91.9, 92.9] | 96.1% [95.1, 97.0] | 96.4% [95.1, 97.3] | 94.0% [90.4, 96.2] |
| 70-79 | 92.2% [91.6, 92.8] | 94.8% [93.8, 95.6] | 95.5% [94.5, 96.4] | 95.4% [93.5, 96.7] |
| 80+ | 85.6% [84.3, 86.7] | 91.2% [89.8, 92.4] | 91.9% [90.4, 93.2] | 92.6% [90.6, 94.1] |
| Overall | 92.8% [92.6, 93.0] | 94.2% [93.6, 94.7] | 94.4% [93.6, 95.0] | 93.7% [92.5, 94.7] |

Table 5 presents the results for the Recovered Cohort when the past PCR-based individualized risk was set to one PCR cluster. Again, the protection was quite similar in all age groups with some decrease in efficacy for the 80+ age category, and quite similar to the results in Table 4. The overall estimated protection of prior SARS-CoV-2 infection for documented recurrent infection was 94·8% (CI: [94·4, 95·1]); hospitalization 94·1% (CI: [91·9, 95·7]); and severe illness 96·4% (CI: [92·5, 98·3]). As there were only 1 death cases in the Recovered Cohort, protection against death was not estimated.

**Table 5:** Protection of prior SARS-CoV-2 infection. Protection of prior SARS-CoV-2 infection for the different age groups adjusted for sex, municipality risk, and past PCR. The overall estimates are based on models without cohort-age interaction. Estimates are not provided for Severe outcomes for the lowest age groups and for Death for all age groups due to very low case numbers in the previously-infected cohorts.

| Age | Positive | Hospitalized | Severe |
| --- | --- | --- | --- |
| 16-39 | 94.5% [94.1, 94.9] | 92.8% [87.3, 95.9] | — |
| 40-49 | 95.1% [94.2, 95.9] | 95.4% [87.7, 98.3] | — |
| 50-59 | 95.2% [94.1, 96.1] | 93.9% [87.1, 97.1] | — |
| 60-69 | 96.1% [94.6, 97.2] | 95.7% [89.6, 98.2] | 96.1% [87.8, 98.7] |
| 70-79 | 97.0% [94.7, 98.3] | 94.1% [86.8, 97.3] | 98.7% [90.5, 99.8] |
| 80+ | 91.4% [85.5, 94.9] | 94.2% [84.5, 97.8] | 94.2% [81.9, 98.1] |
| Overall | 94.8% [94.4, 95.1] | 94.1% [91.9, 95.7] | 96.4% [92.5, 98.3] |

As described above, we assigned the recovered individuals to the middle PCR risk group, so that the estimated protection of a prior infection is compared to unvaccinated individuals having a single PCR cluster in the past. The protection levels afforded by a prior infection compared to unvaccinated persons who had no or 2+ past PCR tests are given in a sensitivity analysis shown in Table S1. In addition, Table S1 presents results of a model without PCR, which can be interpreted as the overall protection of a prior infection. As expected, the protection of a prior infection compared to unvaccinated persons who did not have past PCR tests is estimated to be smaller and compared to those who had 2+ tests is larger. The results when omitting the PCR variable are very similar to the figures in Table 5.

The results for Cohorts 1A and 1B appear in Tables S2 and S3, respectively. The results up to two weeks after the first dose (Cohort 1A) suggest low but statistically significant efficacy. For Cohort 1B that comprises individuals at more than two weeks after the first dose, the efficacy is higher, being 57·7% (CI: [57·1, 58·4]) for documented infection; 69·4% (CI: [67·5, 71·2]) for hospitalization; 65·9% (CI: [63·1, 68·5]) for severe illness; and 62·7% (CI: [58·0, 66·8]) for death. The coefficients of all four models used for analyzing the data appear in Tables S4-S7.

## Discussion

This population-based observational study demonstrates the high efficacy of the BNT162b2 vaccine and prior SARS-CoV-2 infection against both subsequent SARS-CoV-2 infection and other COVID-19–related outcomes. There are a few characteristics that make this study unique. First, it was a nationwide study and thus represented the real-world effectiveness of vaccination and prior infection on the full population. Second, it used individual-level data that enabled, at least to some degree, to mitigate biases caused by selection to get vaccinated, selection to undergo PCR testing, and time-changing level of risk, via adjustment for between-cohort differences in individuals’ characteristics and municipality risk level. Third, the study included follow-up of the population for a period of three months, allowing follow-up of the fully vaccinated cohort over an extended duration. Fourth, this is the first large-scale study that has explored the protection due to prior SARS-CoV-2 infection compared to the Pfizer BNT162b2 vaccine.

There are some limitations to this observational study. One major source of confounding is related to possible population differences between individuals who were vaccinated compare to those who were not. This confounding is partially addressed by controlling for risk factors. Specifically, for each individual we adjusted for sex, age group, number of past PCR tests and the time-dependent environmental exposure. Another major source of potential bias is related to detection of SARS-CoV-2 infection. As apparent from the PCR test counts in Table 3, individuals who are fully vaccinated or were previously infected get tested less often than the unvaccinated cohort. Our results for the outcomes of hospitalization, severe disease, and death do not suffer from this bias and thus are more reliable. The vaccine protection against infection might be biased upward as explained above, nevertheless the remarkable curtailing of the outbreak in Israel which followed the high vaccine uptake by the Israeli population further suggest that the vaccine is efficient in blocking transmission, see Figure 1.

The efficacy estimates of the BNT162b2 vaccine in this study are similar to those reported by previous large-scale studies. For the severe disease outcome, the randomized trial of BNT162b2^1^ reported 89% efficacy for severe disease. A study by the Israeli Ministry of Health using aggregated data^5^ reported 96% efficacy for people as defined in our Cohort 2. A study on data from Israel’s largest HMO^6^ split people as defined in our Cohort 1B and reported an efficacy of 62% and 80% for the third and fourth weeks after the first vaccine, respectively, and of 92% for their Cohort 2. In comparison, our analysis showed efficacy of 66% for Cohort 1B and 94% for Cohort 2. For other outcomes, the estimated vaccine efficacy for Cohort 2 in our study were 93% and 94%, for documented infection and hospitalization, respectively. These estimates are similar to previous studies^5,6^ that estimated efficacy of 92% and 96% for documented infection, and of 87% and 96% for hospitalization. Our findings are based on a longer follow-up and a larger number of event than in the previous individual-level data reports. For example, the analysis of severe cases in the randomized clinical trial is based on only 10 cases, and that of Israel’s largest HMO on 229.^6^ In comparison, the analysis in our study is based on 8,463 cases, including 2,240 cases from Cohort 1 and 319 cases from Cohort 2. On the other hand, the other two studies^1,6^ have the respective advantages of randomization and a detailed matching process which help in bias reduction.

The estimated protection against reinfection in this study is similar to that of the BNT162b2 vaccine. For documented SARS-CoV-2 reinfection, these results are similar to the results obtained in a large study from Qatar of 95% protection,^13^ and suggest higher protection than reported by other previous studies. A large study from Denmark^14^ suggested 80% protection against reinfection. A study on healthcare workers in the United Kingdom^16^ reported that previous infection was associated with an 83% lower risk of infection. These two studies are based on 11,727 and 6,614 previously infected individuals, with 72 and 44 reinfections, respectively. In comparison, the Recovered cohort in our study comprised 187,549 individuals, with 894 reinfections. One possible reason for the differences in the estimated protection against reinfection could be related to detection bias of SARS-CoV-2 infection. However, our estimated high levels of protection against hospitalization and serious disease after reinfection are unlikely to be affected by detection bias, and are reassuring.

An important assumption made here is that rates of infection or hazards are independent of time from vaccination. However, the rate of infection is expected to depend on time from vaccination or on time from first infection. Studying the hazard as a function of time is crucial for understanding waning immunity and for the need for additional booster vaccinations. Follow-up is currently too short to answer time-dependent questions, but this is a crucial and required next step that can be answered using the national Israeli data in the future. The hazard may also depend on calendar time, not only via environmental exposure, but also because of new variants appearing, against which, the vaccine may have different efficacy. During the period over which the data were collected, the COVID-19 variant B.1.1.7 was by far the most prevalent variant, and accounted for most of the documented cases, hence the approximation of a constant hazard is justified. Yet, it is of great importance to repeat this study in other populations in order to estimate the efficacy for other variants and vaccines.

This study suggests that both the BNT162b2 vaccine and prior SARS-CoV-2 infection are effective against both subsequent SARS-CoV-2 infection and other COVID-19–related outcomes. Moreover, the effectiveness seems similar for both cohorts. This puts into question the need to vaccinate recent (up to six month) previously-infected individuals.

## Data Availability

The data used in this study are sensitive and will not be made publicly available.

## Contributors

LF, YG, AH and MM were responsible for study design and for writing of the manuscript. RF, YG, MM, IN, YW, RY, and AZ analyzed the data. YW and AZ were responsible for collecting the data and for data management. LF, YG, AH and MM did the literature survey. LF, RF, YG, MM, and IN developed the mathematical model. YG and YW developed the software for analyzing the data. All authors interpreted the data and reviewed the draft and final versions of the manuscript.

## Ethics statement

The study was approved by the Institutional Review Board of the Sheba Medical Center. Helsinki approval number: SMC-8228-21

## Funding

None

## Competing interests statement

All authors declare no competing interests.

## Data sharing

The data used in this study are sensitive and will not be made publicly available.

## Acknowledgments

The authors would like to thank Guy Katriel for fruitful discussions.

## Web Appendix: The Statistical Model

We define the efficacy of the vaccine in terms of hazard ratios. We use the following constant hazard models to describe the dynamics of an uninfected individual risk over time (calendar time and time from vaccination), where, in the most general model, each cohort has different coefficients:

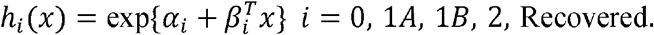

Here *x* indicates a set of risk factors of an individual, including time dependent variables (municipality risk). While the model above is quite general, enabling different coefficients for the different cohorts, our basic model restricts the coefficients of sex, past PCR tests and municipality risk to be equal among the cohorts. Specifically, let

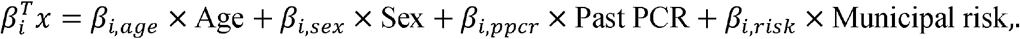

We assume that for *i* = 0, 1*A*, 1*B*, 2, Recovered,

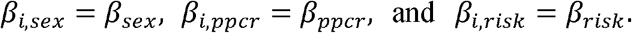

Thus, the effect of sex, past PCR test, and municipal risk on efficacy is multiplicative and identical among cohorts. However, efficacy may vary between different age groups.

The constant hazard assumption implies underlying exponential event-free models for these cohorts, with time-dependent covariates. The analysis can be carried out by performing Poisson regression with offsets for each risk profile. Specifically, consider a group of individuals’ days in Cohort *i* with a certain risk profile *x*_0_ (here the profile also includes time-dependent covariates, so only days satisfying *x*_0_ count). The response variable is ‘case count’ – the number of cases among these individuals’ days, and the exposure is the sum of all at-risk days for individuals with cohort and risk-profile combination (*i, x*_0_). Thus, the model implies

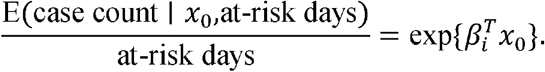

In other words, the daily hazard for an event for an individual in Cohort *i* and risk profile *x*, denoted by *h*_*i,x*_, is 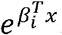. The relative risk for Cohort *i* = 2 with risk profile *x*. is defined as *h*_2,*x*_/*h*_0,*x*_, and the efficacy is defined as 1 − *h*_2,*x*_/*h*_0,*x*_. Under the assumption of equal coefficients for sex, past PCR tests and municipality risk, the relative efficacy depends only on the age group.

Technically, in order to estimate the coefficients in the model, we create a working dataset as follows. For each combination of cohort, age group, sex, municipality risk level, and individualized risk level, we count the number of COVID-19 events and the number of at-risk days. Consider, for example, a 56-year-old male who lives in Tel Aviv, had 1 negative PCR test before December 20, 2020, received his first dose on January 1, 2021, and his second dose on January 23, 2020, and tested positive on February 8, 2021. Assume that the Tel Aviv risk level was category 1 during the period December 20, 2020 to January 20, 2021, category 2 from January 21, 2021 until the end of follow-up on February 8, 2021.

This person contributes:

1. 11 days (Dec-20 to Dec-31) and 0 events to the group: cohort_0/50-60/male/mun_risk=1/past_pcr=1
2. 14 days (Jan-1 to Jan-14) and 0 events to the group: cohort_1A/50-60/male/mun_risk=1/past_pcr=1
3. 6 days (Jan-15 to Jan-20) and 0 events to the group: cohort_1B/50-60/male/mun_risk=1/past_pcr=1
4. 9 days (Jan-21 to Jan-29) and 0 events to the group: cohort_1B/50-60/male/mun_risk=2/past_pcr=1
5. 10 days (Jan-30 to Feb-8) and 1 event to the group: cohort_2/50-60/male/mun_risk=2/past_pcr=1

**Figure S1:**
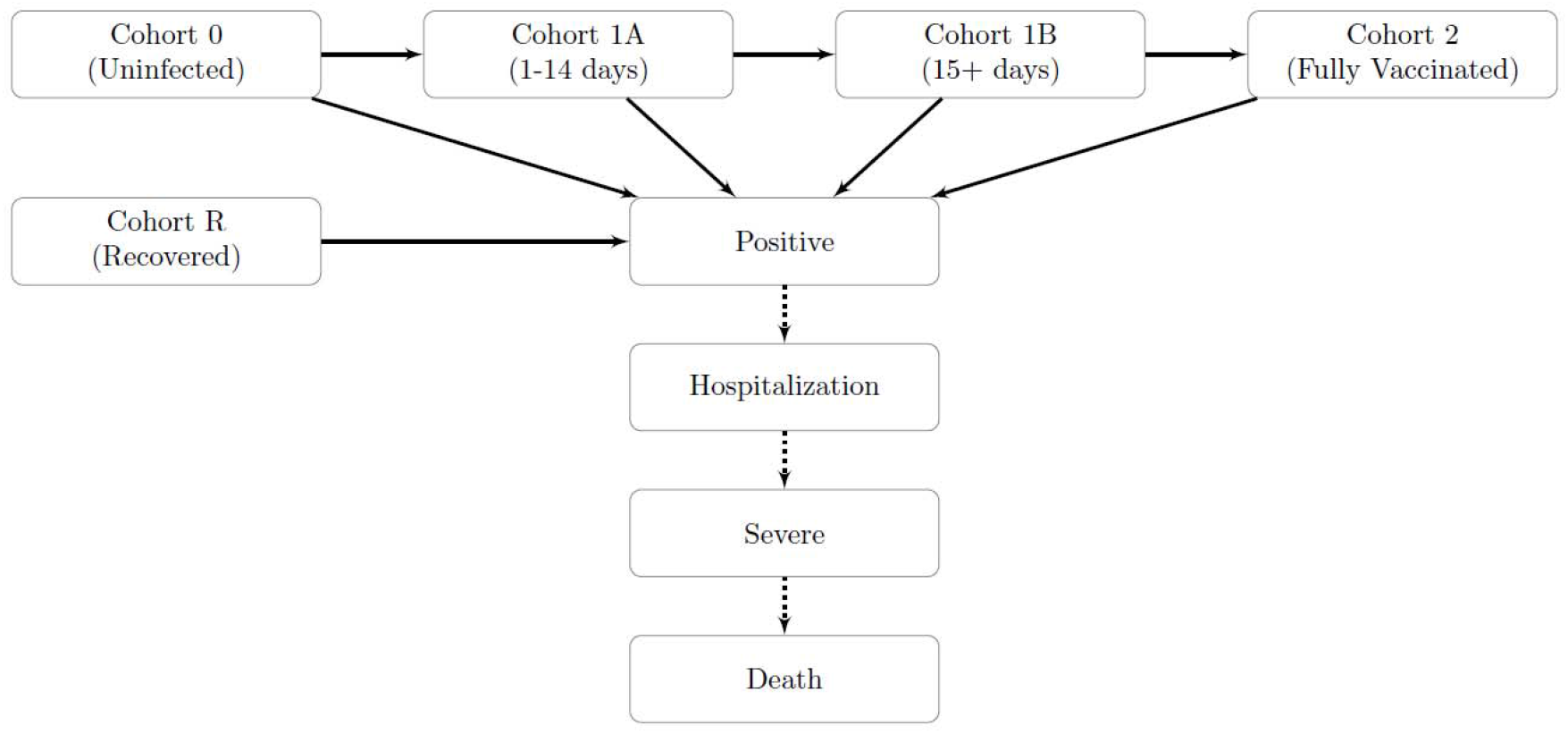
The dynamics of the cohort model. Solid arrows indicate possible transitions between cohorts. Dashed arrows indicate possible disease outcomes.

**Figure S2:**
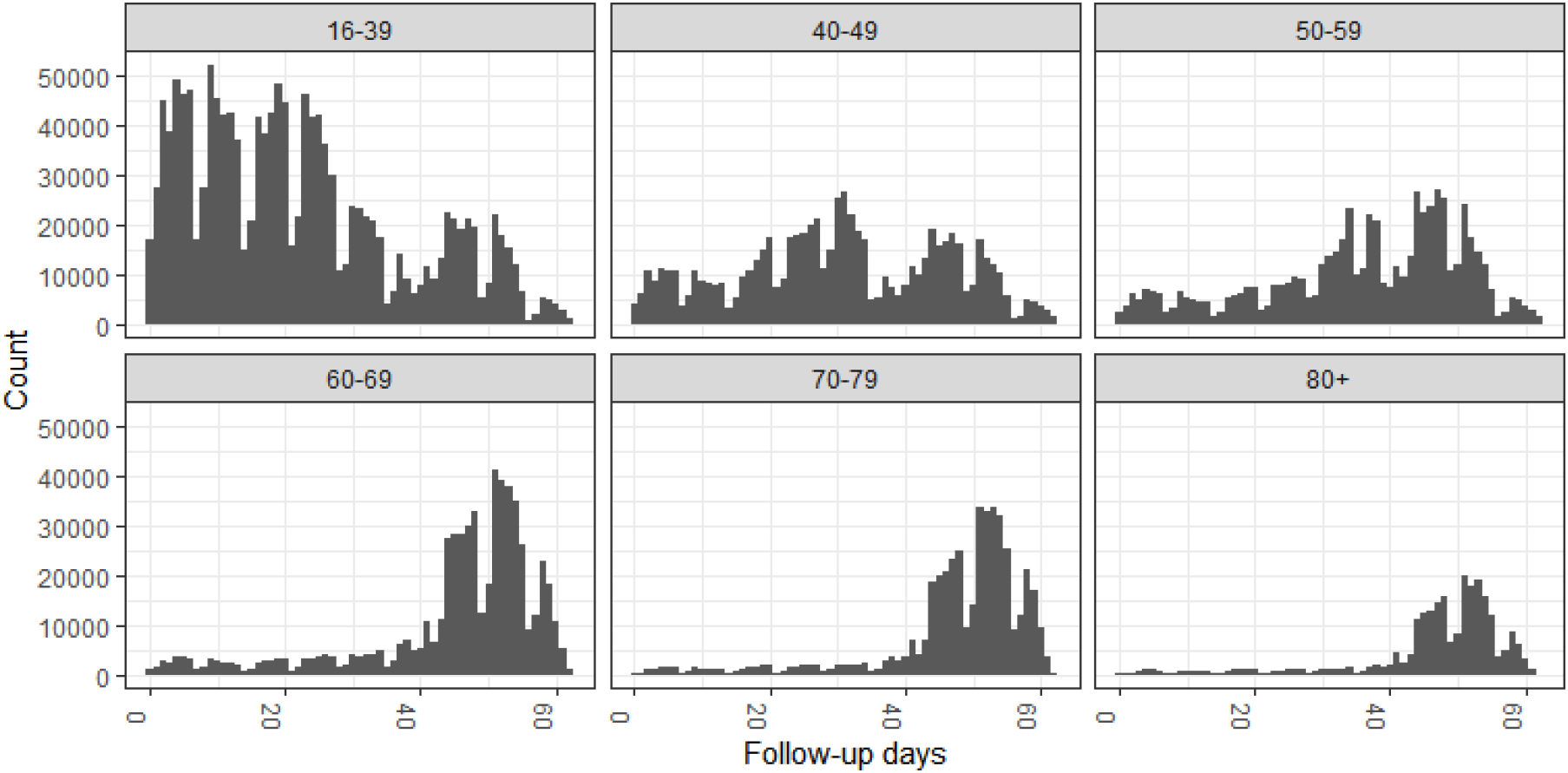
Length of follow-up for Cohort 2. Length of follow-up for Cohort 2 of the fully vaccinated, according to age group. Vaccination became available first to the 60+ age groups and then gradually to younger age groups as can be seen from the follow-up counts.

**Figure S3:**
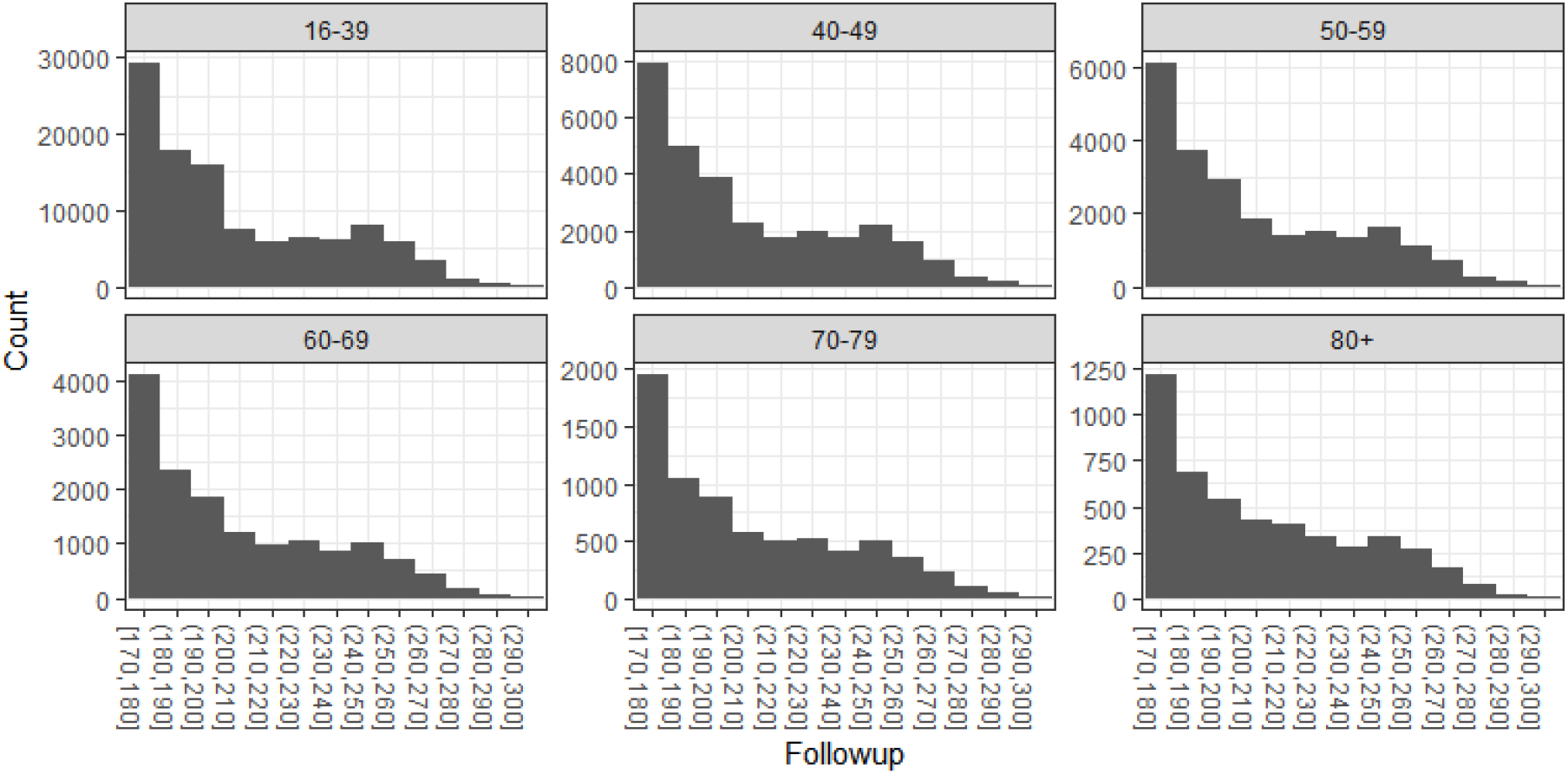
Length of follow-up for the Recovered Cohort. Length of follow-up from first positive PCR test for the Recovered Cohort, according to age group. This cohort included individuals that had a positive PCR test between June 1 and September 30, 2020. Note the sharp decrease in counts as a function of the follow-up. Note that each subfigure has a different scale.

**Figure S4:**
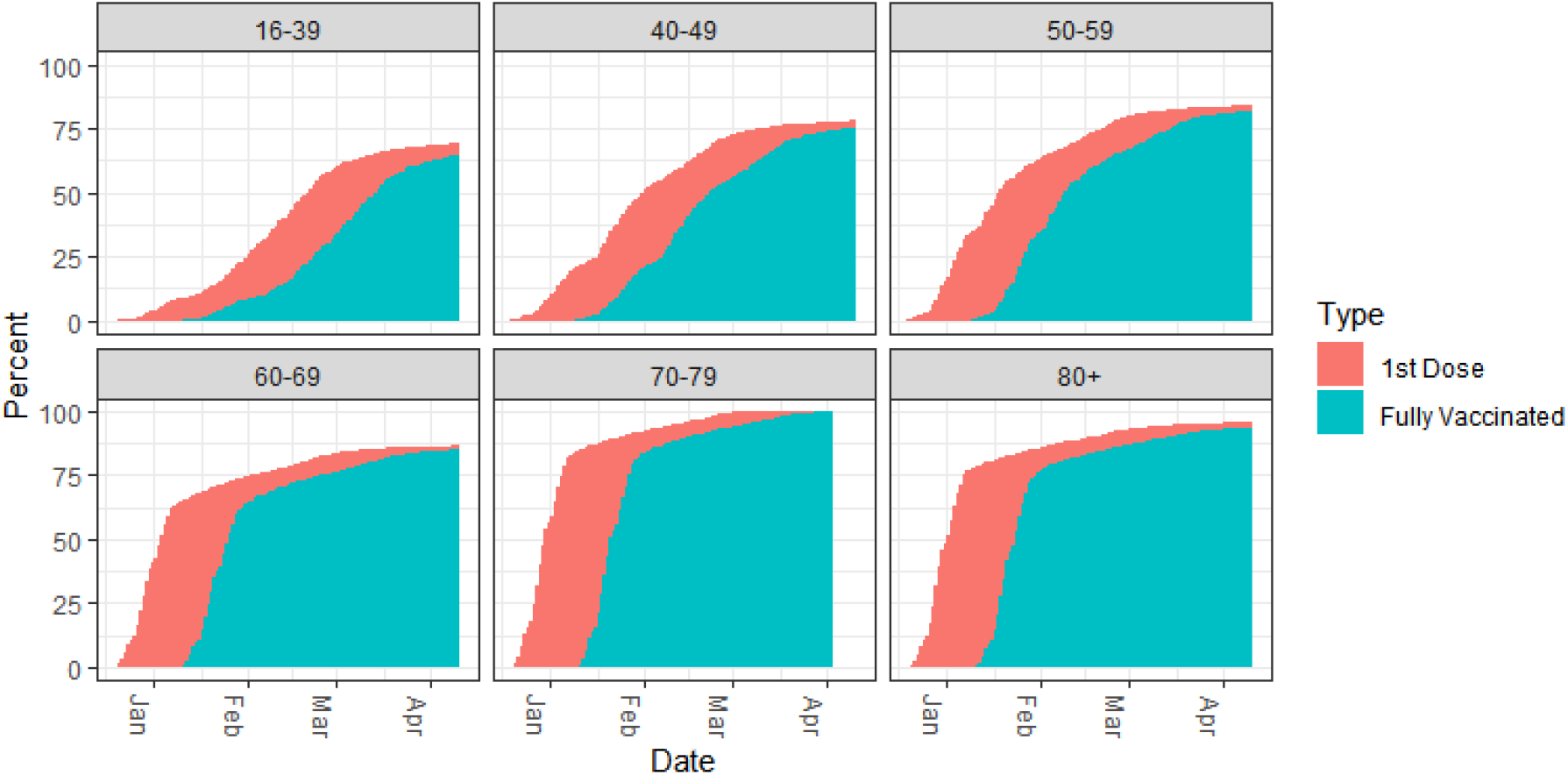
Vaccination by age. Percent of individuals vaccinated with the first and the second dose, by age group. The vaccination initiated in the 60+ age group. See text for details.

**Figure S5:**
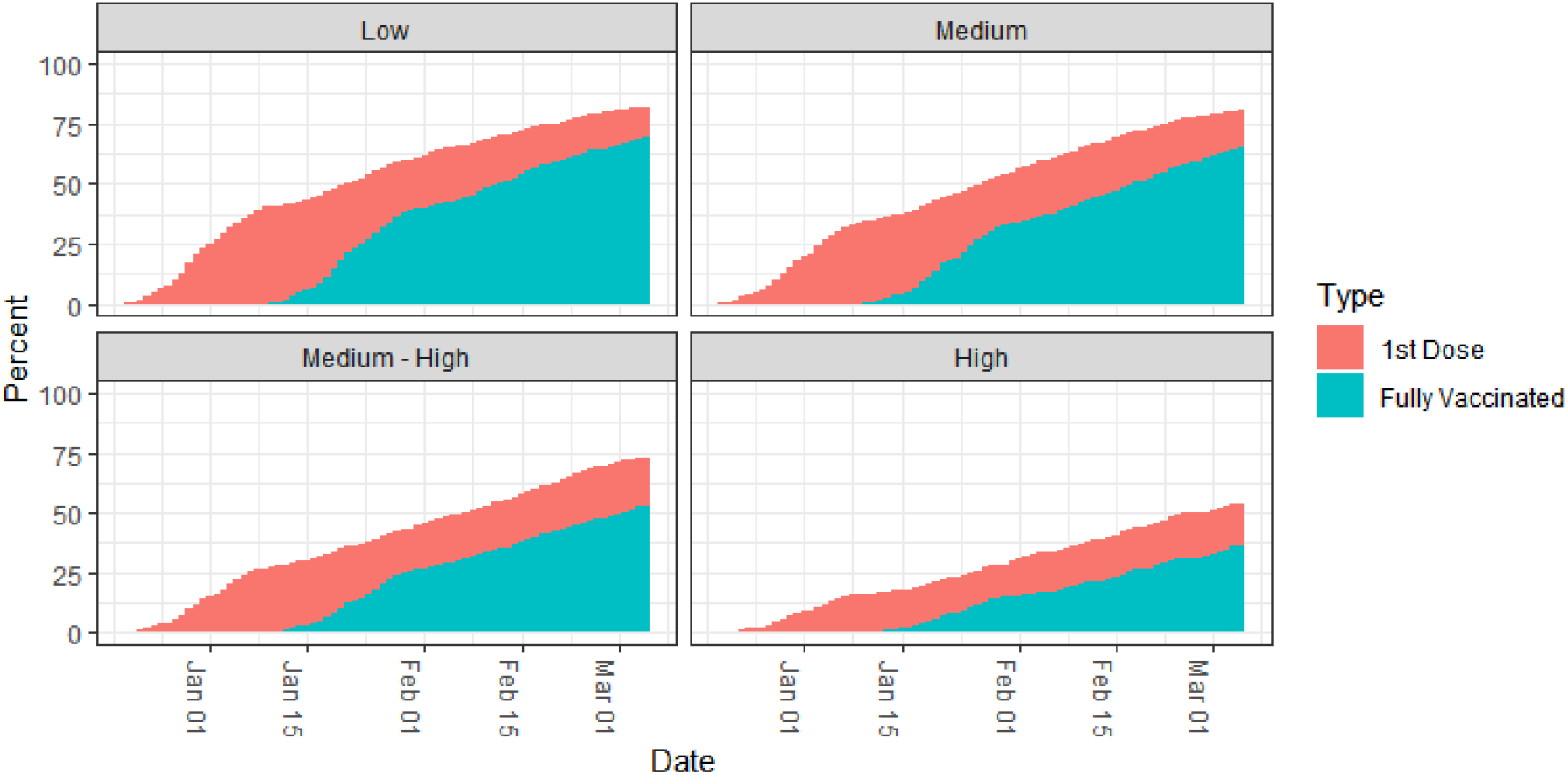
Vaccination by municipality risk. Percent of individuals vaccinated with the first and the second dose, by municipality risk group. The municipality risk was calculated as the median of the daily risk over the research period. Note that there is a negative correlation between vaccine coverage and risk group.

**Figure S6:**
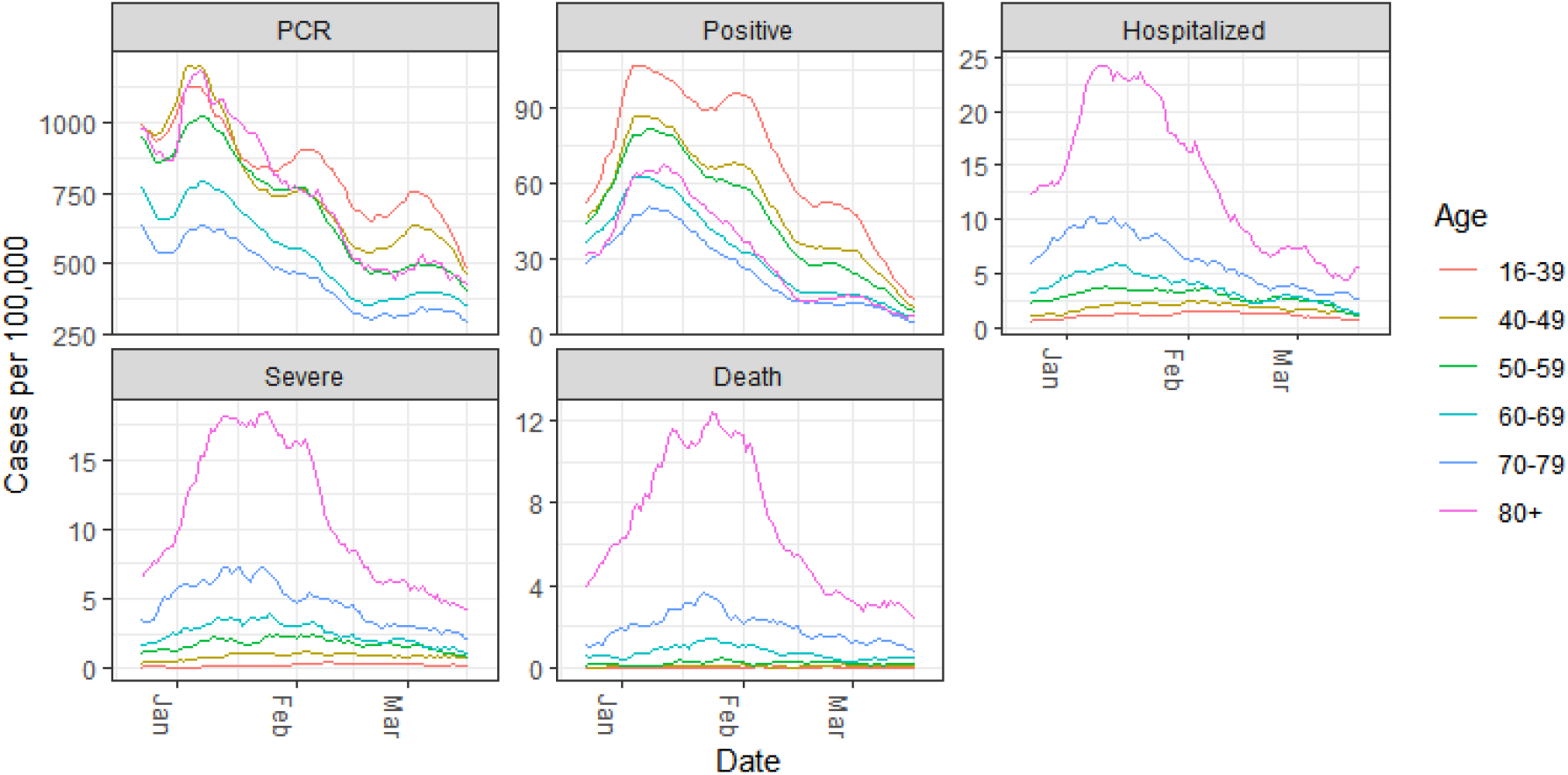
Events over time. Cases per 100,000, smoothed using seven-day moving average for the different age groups and the outcomes: PCR tests, documented infection cases, hospitalized cases, severe cases, and deaths.

**Table S1:** Sensitivity analysis of past PCR on prior SARS-CoV-2 infection. Protection of prior SARS-CoV-2 infection for the different age groups. The model was fitted when the number of PCR clusters is assigned to be 0, 1, 2+, and omitted.

| Analysis | Age | Positive | Hospitalized | Severe |
| --- | --- | --- | --- | --- |
| PCR 0 | 16-39 | 91.9% [91.3, 92.6] | 89.5% [81.5, 94.1] | — |
| PCR 0 | 40-49 | 92.8% [91.4, 93.9] | 93.3% [82.0, 97.5] | — |
| PCR 0 | 50-59 | 92.9% [91.3, 94.3] | 91.0% [81.2, 95.7] | — |
| PCR 0 | 60-69 | 94.2% [92.1, 95.8] | 93.7% [84.8, 97.4] | 94.6% [83.2, 98.3] |
| PCR 0 | 70-79 | 95.6% [92.2, 97.5] | 91.4% [80.7, 96.1] | 98.2% [86.9, 99.7] |
| PCR 0 | 80+ | 87.4% [78.7, 92.5] | 91.6% [77.5, 96.8] | 92.0% [75.1, 97.4] |
| PCR 1 | 16-39 | 94.5% [94.1, 94.9] | 92.8% [87.3, 95.9] | — |
| PCR 1 | 40-49 | 95.1% [94.2, 95.9] | 95.4% [87.7, 98.3] | — |
| PCR 1 | 50-59 | 95.2% [94.1, 96.1] | 93.9% [87.1, 97.1] | — |
| PCR 1 | 60-69 | 96.1% [94.6, 97.2] | 95.7% [89.6, 98.2] | 96.1% [87.8, 98.7] |
| PCR 1 | 70-79 | 97.0% [94.7, 98.3] | 94.1% [86.8, 97.3] | 98.7% [90.5, 99.8] |
| PCR 1 | 80+ | 91.4% [85.5, 94.9] | 94.2% [84.5, 97.8] | 94.2% [81.9, 98.1] |
| PCR 2+ | 16-39 | 95.7% [95.4, 96.1] | 95.4% [91.8, 97.4] | — |
| PCR 2+ | 40-49 | 96.2% [95.5, 96.8] | 97.0% [92.0, 98.9] | — |
| PCR 2+ | 50-59 | 96.3% [95.4, 97.0] | 96.0% [91.7, 98.1] | — |
| PCR 2+ | 60-69 | 97.0% [95.8, 97.8] | 97.2% [93.3, 98.8] | 97.9% [93.3, 99.3] |
| PCR 2+ | 70-79 | 97.7% [95.9, 98.7] | 96.2% [91.5, 98.3] | 99.3% [94.8, 99.9] |
| PCR 2+ | 80+ | 93.3% [88.7, 96.1] | 96.3% [90.0, 98.6] | 96.8% [90.2, 99.0] |
| No PCR | 16-39 | 93.4% [92.9, 93.9] | 91.7% [85.3, 95.3] | — |
| No PCR | 40-49 | 94.0% [92.8, 95.0] | 94.5% [85.4, 98.0] | — |
| No PCR | 50-59 | 94.0% [92.6, 95.2] | 92.6% [84.5, 96.5] | — |
| No PCR | 60-69 | 95.1% [93.2, 96.4] | 94.7% [87.3, 97.8] | 95.5% [86.1, 98.6] |
| No PCR | 70-79 | 96.4% [93.6, 97.9] | 93.2% [84.8, 96.9] | 98.6% [89.8, 99.8] |
| No PCR | 80+ | 90.5% [84.0, 94.4] | 94.0% [84.1, 97.8] | 94.5% [83.0, 98.2] |

**Table S2:** Vaccination efficacy for Cohort 1A. Vaccination efficacy for Cohort 1A adjusted for sex, municipality risk, and past PCR. The overall estimates are based on models without cohort-age interaction. Estimates are not provided for Severe and Death outcomes for the lowest age groups due to very low case numbers in the vaccinated cohorts.

| Age | Positive | Hospitalized | Severe | Death |
| --- | --- | --- | --- | --- |
| 16-39 | 17.7% [16.4, 18.9] | 39.7% [31.0, 47.4] | — | — |
| 40-49 | 17.6% [15.5, 19.6] | 40.7% [31.2, 48.9] | — | — |
| 50-59 | 18.6% [16.3, 20.9] | 44.6% [37.2, 51.1] | — | — |
| 60-69 | 22.4% [19.7, 25.0] | 47.3% [41.2, 52.8] | 49.2% [42.0, 55.6] | 44.7% [28.3, 57.3] |
| 70-79 | 44.0% [41.2, 46.7] | 60.5% [55.9, 64.6] | 62.9% [57.8, 67.3] | 63.6% [55.2, 70.4] |
| 80+ | 17.2% [12.4, 21.7] | 32.6% [26.3, 38.5] | 36.2% [29.2, 42.4] | 40.3% [31.3, 48.1] |
| Overall | 20.6% [19.7, 21.4] | 45.7% [43.1, 48.2] | 49.3% [45.7, 52.7] | 48.5% [42.8, 53.7] |

**Table S3:** Vaccination efficacy for Cohort 1B. Vaccination efficacy for Cohort 1B adjusted for sex, municipality risk, and past PCR. The overall estimates are based on models without cohort-age interaction. Estimates are not provided for Severe and Death outcomes for the lowest age groups due to very low case numbers in the vaccinated cohorts.

| Age | Positive | Hospitalized | Severe | Death |
| --- | --- | --- | --- | --- |
| 16-39 | 67.3% [66.4, 68.1] | 82.4% [77.0, 86.6] | — | — |
| 40-49 | 55.1% [53.6, 56.6] | 82.8% [77.3, 87.0] | — | — |
| 50-59 | 50.3% [48.5, 52.0] | 80.7% [76.3, 84.3] | — | — |
| 60-69 | 48.6% [46.6, 50.6] | 70.0% [65.6, 73.8] | 71.4% [66.3, 75.8] | 63.3% [50.5, 72.7] |
| 70-79 | 56.2% [53.9, 58.5] | 70.4% [66.6, 73.7] | 72.6% [68.5, 76.1] | 72.1% [65.1, 77.7] |
| 80+ | 36.6% [32.6, 40.3] | 54.1% [49.2, 58.6] | 55.8% [50.4, 60.6] | 56.6% [49.3, 62.8] |
| Overall | 57.7% [57.1, 58.4] | 69.4% [67.5, 71.2] | 65.9% [63.1, 68.5] | 62.7% [58.0, 66.8] |

**Table S4:**
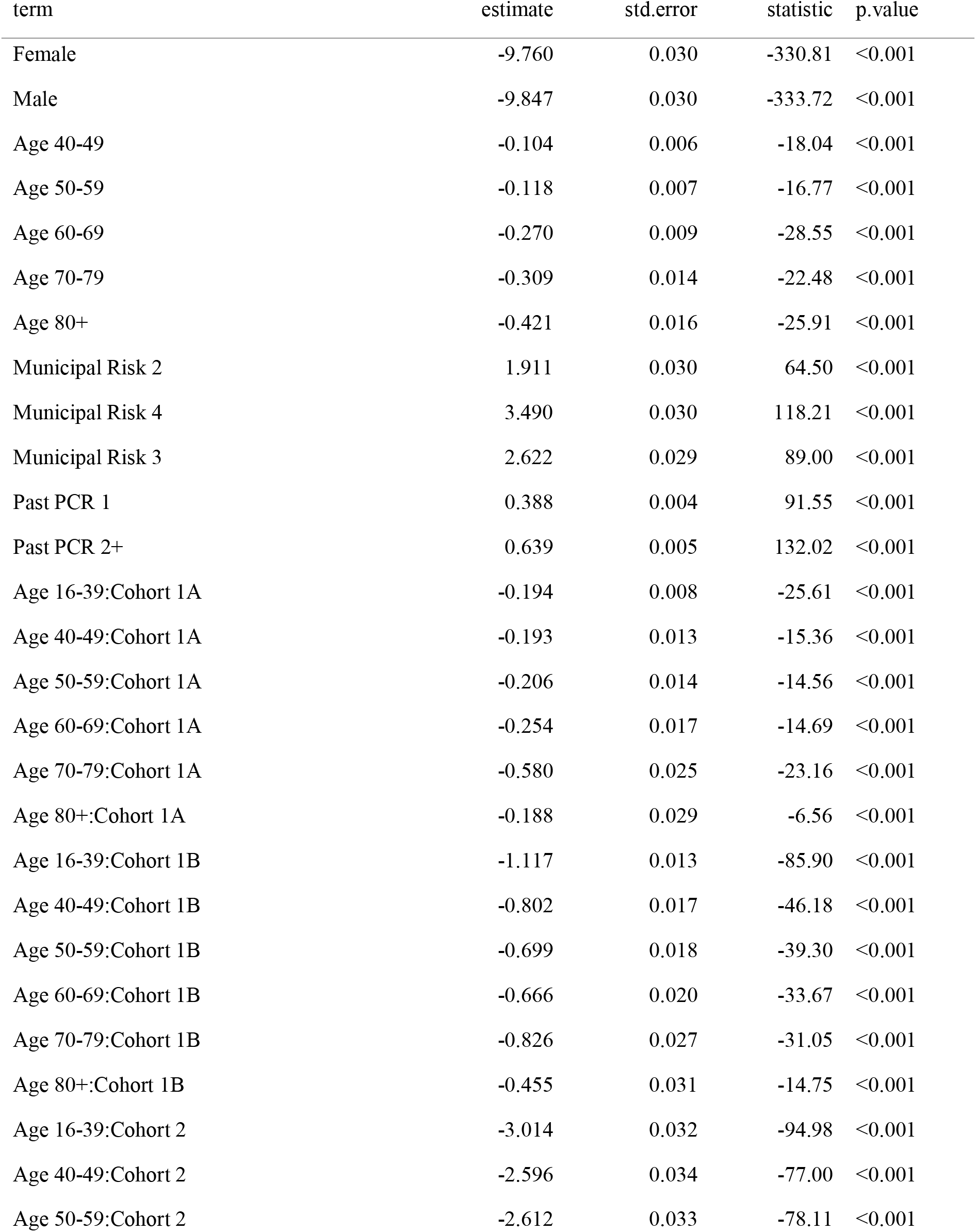
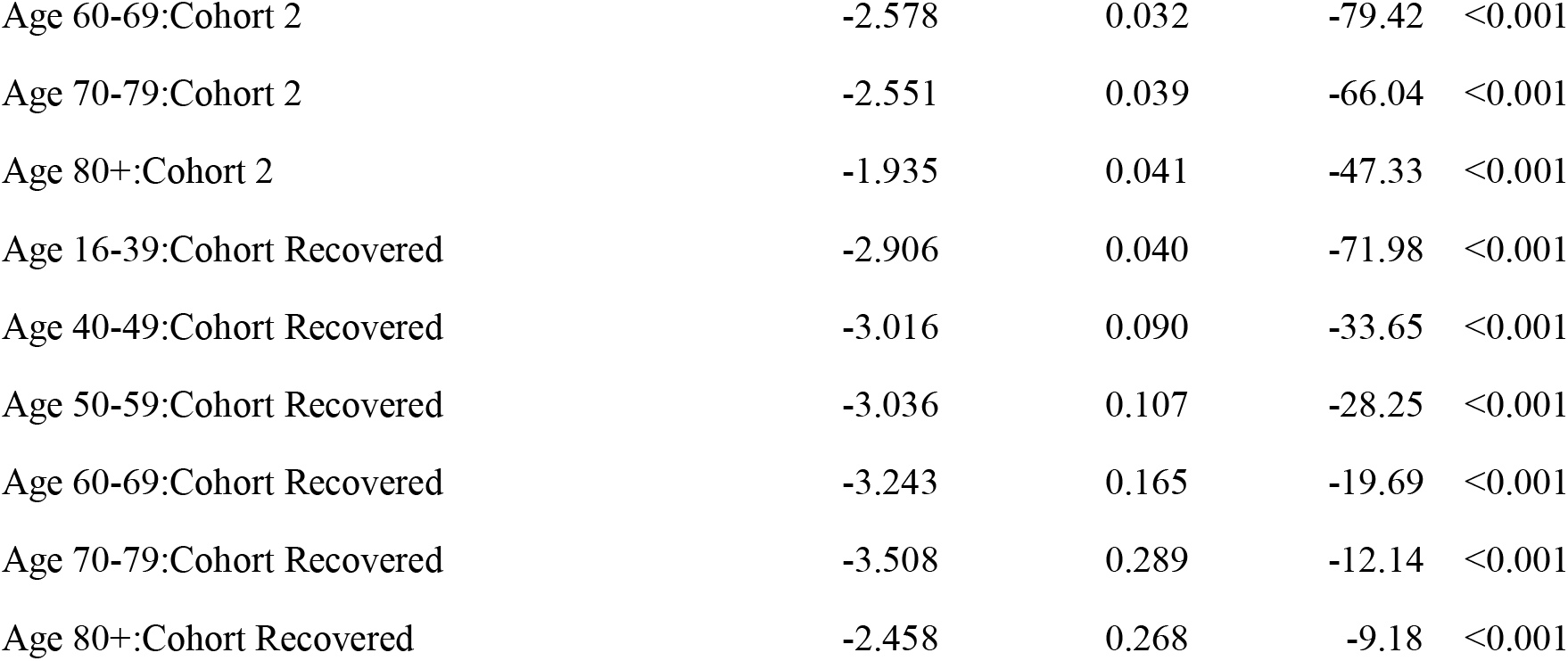
Model coefficients for the documented infection outcome.

**Table S5:** Model coefficients for the hospitalization outcome.

| term | estimate | std.error | statistic | p.value |
| --- | --- | --- | --- | --- |
| Female | -13.605 | 0.114 | -119.280 | <0.001 |
| Male | -13.469 | 0.114 | -118.210 | <0.001 |
| Age 40-49 | 0.733 | 0.034 | 21.451 | <0.001 |
| Age 50-59 | 1.332 | 0.033 | 40.784 | <0.001 |
| Age 60-69 | 1.829 | 0.033 | 55.740 | <0.001 |
| Age 70-79 | 2.525 | 0.033 | 75.708 | <0.001 |
| Age 80+ | 2.940 | 0.032 | 92.933 | <0.001 |
| Municipal Risk 2 | 1.488 | 0.113 | 13.140 | <0.001 |
| Municipal Risk 4 | 2.902 | 0.113 | 25.636 | <0.001 |
| Municipal Risk 3 | 2.219 | 0.112 | 19.726 | <0.001 |
| Past PCR 1 | 0.377 | 0.021 | 18.111 | <0.001 |
| Past PCR 2+ | 0.815 | 0.021 | 38.294 | <0.001 |
| Age 16-39:Cohort 1A | -0.507 | 0.069 | -7.349 | <0.001 |
| Age 40-49:Cohort 1A | -0.522 | 0.076 | -6.901 | <0.001 |
| Age 50-59:Cohort 1A | -0.590 | 0.064 | -9.219 | <0.001 |
| Age 60-69:Cohort 1A | -0.641 | 0.056 | -11.402 | <0.001 |
| Age 70-79:Cohort 1A | -0.928 | 0.056 | -16.705 | <0.001 |
| Age 80+:Cohort 1A | -0.395 | 0.046 | -8.559 | <0.001 |
| Age 16-39:Cohort 1B | -1.740 | 0.138 | -12.638 | <0.001 |
| Age 40-49:Cohort 1B | -1.760 | 0.141 | -12.451 | <0.001 |
| Age 50-59:Cohort 1B | -1.645 | 0.105 | -15.637 | <0.001 |
| Age 60-69:Cohort 1B | -1.204 | 0.069 | -17.328 | <0.001 |
| Age 70-79:Cohort 1B | -1.216 | 0.060 | -20.128 | <0.001 |
| Age 80+:Cohort 1B | -0.779 | 0.052 | -14.943 | <0.001 |
| Age 16-39:Cohort 2 | -3.353 | 0.289 | -11.582 | <0.001 |
| Age 40-49:Cohort 2 | -2.882 | 0.198 | -14.546 | <0.001 |
| Age 50-59:Cohort 2 | -3.005 | 0.153 | -19.641 | <0.001 |
| Age 60-69:Cohort 2 | -3.254 | 0.123 | -26.432 | <0.001 |
| Age 70-79:Cohort 2 | -2.949 | 0.089 | -33.302 | <0.001 |
| Age 80+:Cohort 2 | -2.425 | 0.074 | -32.629 | <0.001 |
| Age 16-39:Cohort Recovered | -2.635 | 0.290 | -9.093 | <0.001 |
| Age 40-49:Cohort Recovered | -3.074 | 0.501 | -6.137 | <0.001 |
| Age 50-59:Cohort Recovered | -2.790 | 0.379 | -7.359 | <0.001 |
| Age 60-69:Cohort Recovered | -3.138 | 0.448 | -7.000 | <0.001 |
| Age 70-79:Cohort Recovered | -2.826 | 0.409 | -6.903 | <0.001 |
| Age 80+:Cohort Recovered | -2.849 | 0.501 | -5.689 | <0.001 |

**Table S6:** Model coefficients for the severe disease outcome. Estimates are not provided for the lowest age groups due to very low event counts in the vaccinated cohorts.

| term | estimate | std.error | statistic | p.value |
| --- | --- | --- | --- | --- |
| Female | -12.258 | 0.177 | -69.170 | <0.001 |
| Male | -11.829 | 0.177 | -66.859 | <0.001 |
| Age 70-79 | 0.807 | 0.043 | 18.716 | <0.001 |
| Age 80+ | 1.261 | 0.041 | 30.411 | <0.001 |
| Municipal Risk 2 | 1.400 | 0.176 | 7.944 | <0.001 |
| Municipal Risk 4 | 2.902 | 0.176 | 16.445 | <0.001 |
| Municipal Risk 3 | 2.158 | 0.175 | 12.321 | <0.001 |
| Past PCR 1 | 0.321 | 0.035 | 9.285 | <0.001 |
| Past PCR 2+ | 0.929 | 0.032 | 29.362 | <0.001 |
| Age 60-69:Cohort 1A | -0.678 | 0.068 | -9.914 | <0.001 |
| Age 70-79:Cohort 1A | -0.991 | 0.065 | -15.281 | <0.001 |
| Age 80+:Cohort 1A | -0.449 | 0.053 | -8.513 | <0.001 |
| Age 60-69:Cohort 1B | -1.253 | 0.085 | -14.751 | <0.001 |
| Age 70-79:Cohort 1B | -1.293 | 0.071 | -18.248 | <0.001 |
| Age 80+:Cohort 1B | -0.817 | 0.059 | -13.808 | <0.001 |
| Age 60-69:Cohort 2 | -3.313 | 0.152 | -21.748 | <0.001 |
| Age 70-79:Cohort 2 | -3.104 | 0.109 | -28.587 | <0.001 |
| Age 80+:Cohort 2 | -2.515 | 0.087 | -28.962 | <0.001 |
| Age 60-69:Cohort Recovered | -3.237 | 0.579 | -5.593 | <0.001 |
| Age 70-79:Cohort Recovered | -4.314 | 1.001 | -4.311 | <0.001 |
| Age 80+:Cohort Recovered | -2.845 | 0.579 | -4.918 | <0.001 |

**Table S7:** Model coefficients for the death outcome. Estimates are not provided for the lowest age groups and the Recovered cohort due to very low event counts.

| term | estimate | std.error | statistic | p.value |
| --- | --- | --- | --- | --- |
| Female | -13.685 | 0.259 | -52.938 | <0.001 |
| Male | -13.096 | 0.258 | -50.798 | <0.001 |
| Age 70-79 | 1.227 | 0.079 | 15.612 | <0.001 |
| Age 80+ | 2.072 | 0.072 | 28.798 | <0.001 |
| Municipal Risk 2 | 1.284 | 0.253 | 5.066 | <0.001 |
| Municipal Risk 4 | 2.760 | 0.254 | 10.864 | <0.001 |
| Municipal Risk 3 | 2.023 | 0.252 | 8.038 | <0.001 |
| Past PCR 1 | 0.393 | 0.055 | 7.198 | <0.001 |
| Past PCR 2+ | 1.202 | 0.046 | 25.975 | <0.001 |
| Age 60-69:Cohort 1A | -0.592 | 0.132 | -4.489 | <0.001 |
| Age 70-79:Cohort 1A | -1.010 | 0.106 | -9.534 | <0.001 |
| Age 80+:Cohort 1A | -0.515 | 0.071 | -7.218 | <0.001 |
| Age 60-69:Cohort 1B | -1.002 | 0.152 | -6.593 | <0.001 |
| Age 70-79:Cohort 1B | -1.277 | 0.114 | -11.191 | <0.001 |
| Age 80+:Cohort 1B | -0.834 | 0.079 | -10.606 | <0.001 |
| Age 60-69:Cohort 2 | -2.811 | 0.238 | -11.823 | <0.001 |
| Age 70-79:Cohort 2 | -3.081 | 0.174 | -17.743 | <0.001 |
| Age 80+:Cohort 2 | -2.599 | 0.119 | -21.889 | <0.001 |

## Notes

### Competing Interest Statement

The authors have declared no competing interest.

